# Revisiting the association of pesticide exposure and Parkinson’s disease: systematic review and meta-analysis

**DOI:** 10.1101/2025.08.19.25333681

**Authors:** Pedro Henrique Passos da Silva, Jadson Silva Abreu, Rafael Sidônio Gibson Gomes, Hemengella Karyne Alves Oliveira, Artur F. S. Schuh, Ignacio F. Mata, Bruno Lopes Santos-Lobato

## Abstract

**Background:** The association between pesticide exposure and Parkinson’s disease (PD) is substantial, but heterogeneity in methodology and lack of categorization according to the type of exposure and pesticide classes in previous meta-analyses impair the interpretation of data. This study aims to update evidence of the association between pesticide exposure and PD.

**Methods:** We conducted a systematic review and meta-analysis of studies investigating associations between pesticide exposure and PD according to the type of pesticide exposure and pesticide class. We searched PubMed, EMBASE, and Web of Science until July 2024. Reviewers screened titles and abstracts. Afterward, reviewers reanalyzed the selection criteria and extracted the data based on the full paper. Meta-analyses were conducted to assess the association between pesticide exposure and PD.

**Results:** A total of 124 studies were eligible. There is a lack of diversity in the populations represented and a high variability in methodology among the included studies. Considering only studies with any type of exposure, we found a positive association of PD with any pesticide class and herbicides. Occupational exposure was associated with PD for all pesticide classes except for fungicides. Exclusive household pesticide exposure was also associated with PD.

**Conclusion:** Pesticide exposure remains a significant environmental risk factor for the development of PD, regardless of the type of exposure. Herbicides are the pesticide class with the most substantial evidence of association with the disease. Further studies with new methods of pesticide exposure measurement, innovative design studies, and the inclusion of underrepresented populations are still needed.

## 1 INTRODUCTION

The growing incidence of Parkinson’s disease (PD) worldwide in the last decades ^1^ has led to a renewed effort to find an association between environmental risk factors associated with the condition, with a focus on pesticides. Since the first descriptions of early-onset PD in two Italians with extensive pesticide use in 1986 ^2^ and two Brazilian rural workers caused by chronic exposure to the herbicide maneb in 1988 ^3^, many other epidemiological studies have explored the association, which has been confirmed by most of these works. Previous systematic reviews with meta-analyses aggregated these data, showing that pesticide exposure increases the risk of developing PD ^4–14^.

However, the methodological heterogeneity of these studies has recently been highlighted ^15^, which may impair the generalizability of the results and weaken the level of evidence supporting the association between pesticide exposure and PD. Additionally, most previous systematic reviews did not differentiate between studies based on the type of exposure (occupational or household) or pesticide classes (insecticides, herbicides, or fungicides). To update and improve the evidence of the association, we conducted a systematic review and meta-analysis of epidemiological studies on pesticide exposure and PD.

## 2 MATERIAL AND METHODS

### 2.1 Design

We conducted a systematic review with a meta-analysis of original observational studies (cross-sectional, case-control, cohort) investigating associations between pesticide exposure and PD, published in English, Spanish, and Portuguese. We excluded studies that (1) assessed other parkinsonian conditions (such as atypical parkinsonism and dementia with Lewy bodies); (2) were conducted on animal models or in vitro; (3) review studies; (4) preprint articles; and (6) duplicated articles. We followed the Preferred Reporting Items for Systematic Reviewers and Meta-Analyses (PRISMA) 2020 guidelines to elaborate this review. The protocol was registered in the PROSPERO database (2024 CRD42024585463).

### 2.2 Search strategy and selection process

Two independent reviewers searched the literature in three databases (PubMed, EMBASE, and Web of Science) from inception to July 2024. The search strategy included terms for pesticide exposure and PD.

All studies were imported into the Rayyan-Intelligent Systematic Review software ^16^, which is used for recording decisions. In the first round, two reviewers screened articles, blindly assessing titles and abstracts based on the inclusion and exclusion criteria. Disagreements were resolved by consensus with the assistance of a third reviewer.

For the second round, selected articles were randomly assigned to two reviewers who evaluated the full texts for reanalyzing selection criteria and extracted the data using a structured online spreadsheet, in which the following items were reported: (1) first author’s name; (2) year of publication; (3) geographical location of the studied population; (4) sample size in people with PD and controls; (5) sex and age at evaluation in people with PD and controls; (6) rural/urban origin of the studied population; (7) type of pesticide exposure (occupational/household/any exposure); (8) pesticide class evaluated (insecticide/herbicide/fungicide/any class); (9) number of people with PD exposed and non-exposed to pesticides; (10) number of controls exposed and non-exposed to pesticides. Disagreements were resolved by reaching a consensus between reviewers.

When extracting data on the number of participants (with PD or controls) exposed or not exposed to pesticides in each study, we categorized it according to the authors’ description of pesticide exposure as “occupational” or “household”; if categorization was not possible due to the lack of information, it was labeled as “any exposure.” The same categorization method based on the authors’ description was used for the pesticide class (“insecticide,” “herbicide,” “fungicide”; if categorization was not possible, “any class”). When the study examined multiple pesticides, we extracted data from the most commonly used products with positive associations to represent pesticide class exposure.

### 2.3 Quality assessment

The quality of each selected study was evaluated using an instrument for exposure/outcome case-control studies in meta-analyses, the Newcastle-Ottawa Quality Assessment Scale (NOS) ^17^. This tool is based on eight items that assess the following themes: case definition, representativeness of cases, selection of controls, the definition of controls, comparability of cases and controls, ascertainment of exposure, a similar method of ascertainment for cases and controls, and non-response rate. Points were attributed to each item, and a score ranging from 0 to 9 was generated for each study. Scores 0-2 on the NOS indicate poor-quality studies, 3-5 indicate fair quality, and 6-9 indicate high-quality studies.

### 2.4 Data analysis

To estimate the association between pesticide exposure and PD, meta-analyses of original case-control studies were performed using R software version 4.0.4 (R Core Team 2018) with the package *metafor*. We used the random-effects model with a continuity correction of 0.5 for studies with individuals without pesticide exposure in the PD or control group. The odds ratio (OR) of developing PD among individuals with pesticide exposure was calculated using the restricted maximum likelihood (REML) estimator.

Egger’s test was used to investigate publication bias. We also used Higgin’s and Thompson’s I^2^ statistics to analyze heterogeneity between studies.

To identify potential sources of heterogeneity in the meta-analysis, we conducted univariate meta-regression analyses among studies included in the meta-analysis that showed significant associations, using a mixed-effects model. This analysis considered study quality and methodological factors, including rural/urban setting, number of participants, study origin (global subregion), method of exposure evaluation (questionnaire, geocoded, or pesticide measurement), and publication year.

Also, we performed sensitivity analyses to explore: (I) the association of pesticide exposures with PD without studies from two leading research groups (Ritz group - USA - 18 studies; Elbaz group - France - 7 studies), aiming to reduce the overlap effect of including the same participant in different publications from the group, and (II) study quality through two subgroups (NOS < 6, poor and fair quality; NOS ≥ 6, high quality).

## 3 RESULTS

### 3.1 Characteristics of included studies

We identified 5,686 publications, of which 207 met the selection criteria for the first round. After the second round, 124 studies were eligible. The PRISMA 2020 flowchart (Figure 1) summarizes the step-by-step selection process.

**Figure 1.**
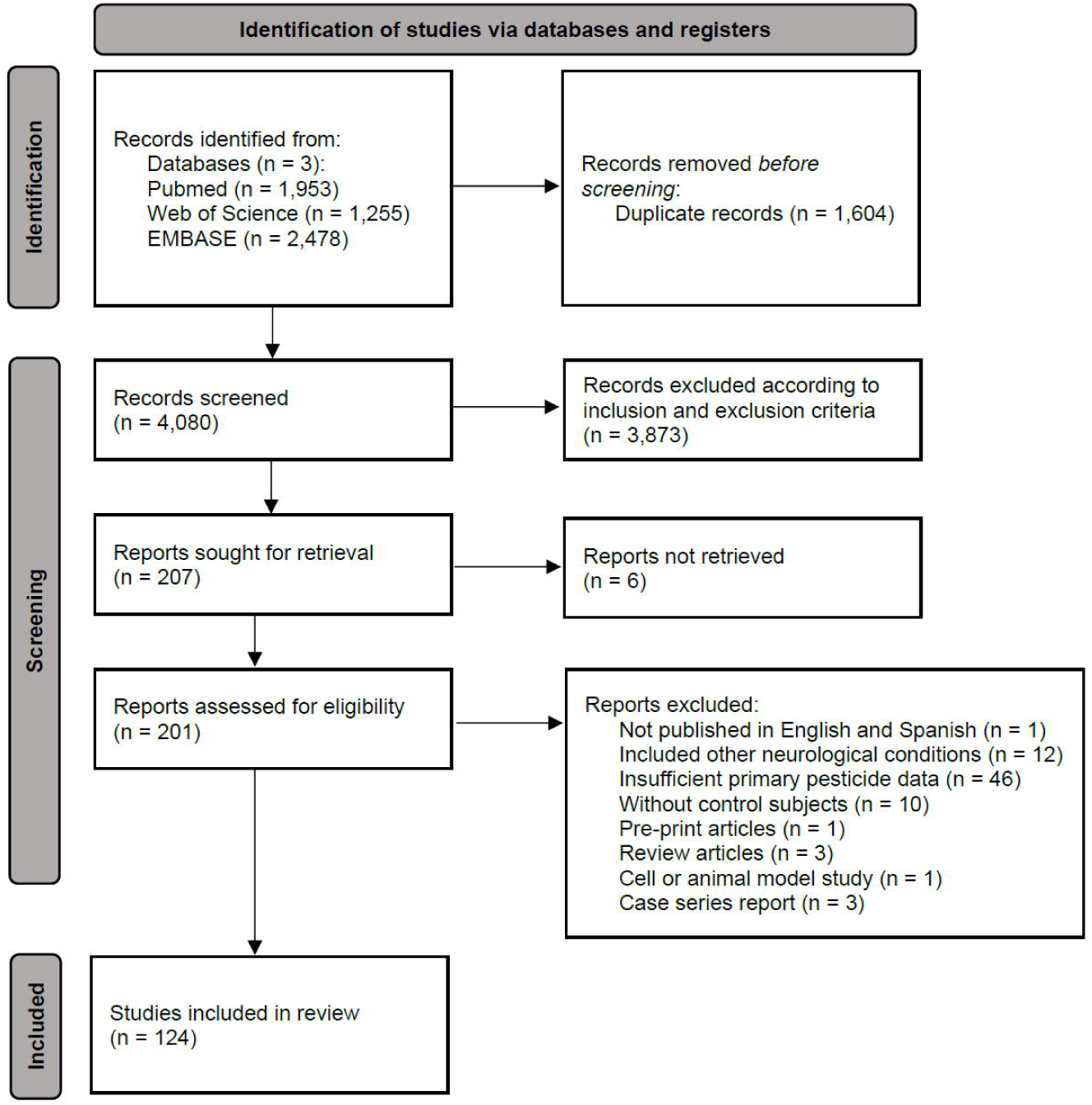
PRISMA 2020 flowchart.

Studies from 35 different countries were included. Most eligible studies were from North America (n = 55), with 48 studies from the USA. From Europe, 35 studies were selected, nine from France. Participation from other regions (Asia, n = 19; Central and South America, n = 6; Oceania, n = 6; Africa, n = 2) was more limited (Figure 2). According to the number of participants per region, the majority of recruited individuals were from North America, with Central and South America, and Africa contributing the fewest (Table 1). Regarding the sample size, only four studies enrolled more than 1,000 people with PD. The quality assessment identified that most studies had fair quality (NOS 3-5; n = 85), with 21 studies classified as high quality (NOS ≥ 6).

**Figure 2.**
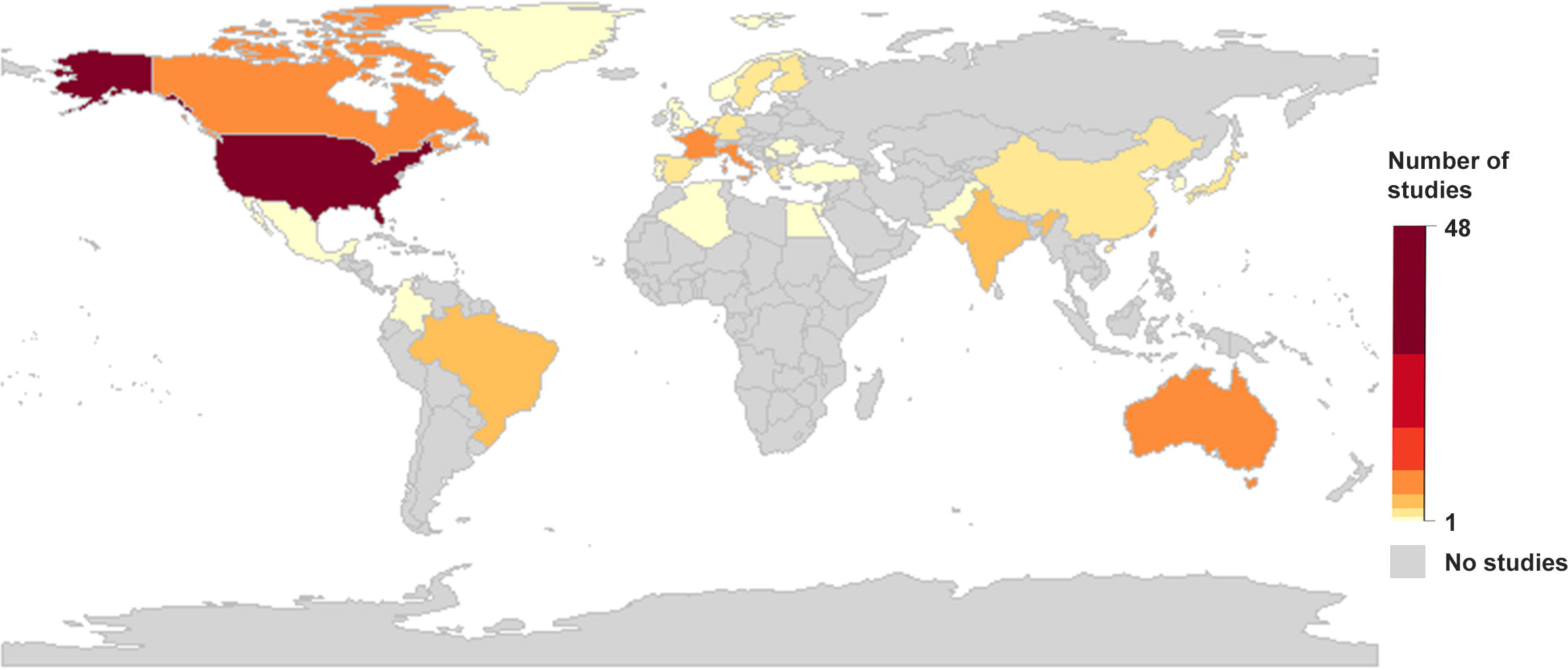
Selected studies count by countries/regions.

**Table 1.** Number of participants from selected studies per global subregion.

| **Global subregion** | **n Studies** | **n People with PD** | **n Male People with PD** | **n Female People with PD** | **n Controls** | **n Male Controls** | **n Female Controls** |
| --- | --- | --- | --- | --- | --- | --- | --- |
| North America | 55 | 15,210 | 8,180* | 5,190* | 314,302 | 53,942* | 36,498* |
| Europe | 35 | 9,624 | 5,368 | 3,969 | 160,897 | 87,421 | 71,973 |
| Asia | 19 | 3,852 | 2,128 | 1,576 | 50,653 | 34,139 | 15,821 |
| Central and South America | 6 | 1,163 | 493* | 416* | 1,119 | 401* | 549* |
| Oceania | 6 | 2,086 | 1,117* | 634* | 2,004 | 924* | 759* |
| Africa | 2 | 737 | 427 | 310 | 767 | 437 | 330 |
| Total | 124<sup>§</sup> | 32,825 | 17,785* | 12,176* | 529,897 | 177,419* | 126,019* |

Abbreviations: n, total number of participants (counts) included in studies; PD, Parkinson's disease.
§ One study [e123] could not be categorized by subregion due to a global recruitment strategy (genetic testing company consumers)
\* Studies that did not describe the number of male and female participants: North America – 7 studies; Central and South America – 1 study; Oceania – 1 study

Only 18 studies were population-based (usually rural communities), and 38 studies recruited participants exclusively from rural regions. According to the study design, only two of the 124 studies were longitudinal cohorts, while the others were case-control or cross-sectional studies. A total of 22 studies performed at least one sensitivity analysis. Movement disorders specialists confirmed the diagnosis of PD in 37 studies, general neurologists confirmed it in 67 studies, and the remaining 20 studies employed different strategies to confirm the PD diagnosis (non-neurologist clinicians, self-reported diagnosis, record linkage, students). There was a slight predominance of males among the recruited participants, with males comprising 58.5% of individuals with PD and 53.4% of the control group. The mean age at evaluation of people with PD and controls were 67.0 and 66.3 years, respectively. The mean age at disease onset was 61.5 years.

Regarding pesticide data, exposure was quantified through environmental questionnaires in 102 studies, exposure estimates (geocoded data or job exposure matrix) in 14 studies, and exposure was based on measuring organochlorines in blood in seven studies. In one study, the data were based on a poisoning registry. Only 11 studies based on questionnaires tested the reliability of the instrument.

Exclusive occupational pesticide exposure was analyzed in 34 studies, exclusive household pesticide exposure in only five studies, and most evaluated both exposures (combined – any exposure, n = 58; segregated - occupational and household, n = 27). Pesticide classes were categorized in 61 studies, with the specific pesticides most frequently explored being paraquat (n = 16) and maneb (n = 8). Most studies (n = 82) defined pesticide exposure as any contact during participants’ lifetime, and 18 studies included definitions of high exposure for analyses, usually based on median values of duration and frequency of pesticide usage. We noted a marked variability in methodological procedures among studies on pesticide exposure measurement and analyses.

### 3.2 Association between any type of pesticide exposure and Parkinson’s disease

Considering any type of pesticide exposure (occupational or household), the meta-analysis showed that the use of any pesticide class (n = 44 studies) during lifetime increased the association with PD (OR = 1.80; 95% CI = 1.51 to 2.14) (Table 2), with high heterogeneity among studies (I^2^ statistics = 83.4%). For herbicides (n = 11), the meta-analyses showed that the use of this pesticide class during a lifetime increased the association with PD (OR = 1.58; 95% CI = 1.26 to 1.97) (Table 2), with high heterogeneity (I^2^ statistics = 72.4%). For fungicides (n = 4; number of participants = 2,649), the meta-analysis showed no association with PD (OR = 1.45; 95% CI = 0.70 to 2.99) (Table 2), but for insecticides (n = 12), the meta-analyses showed a trend of increasing the association with PD (OR = 1.43; 95% CI = 1.00 to 2.05, p = 0.0524) (Table 2), both with high heterogeneity (fungicides: I^2^ statistics = 78.5%; insecticides: I^2^ statistics = 87.7%). According to Egger’s test, no publication bias was detected among the studies.

**Table 2.** Meta-analysis of the association between pesticide exposure and Parkinson’s disease.

| Exposure | Pesticide class | K | n | Random-effects model |  |  | Heterogeneity<br>I<sup>2</sup> | Egger's test |
| --- | --- | --- | --- | --- | --- | --- | --- | --- |
|  |  |  |  | OR | 95% CI | p-value |  |  |
| Any type | Any pesticides | 44 | 171,767 | **1.80** | **1.51-2.14** | **< 0.0001** | 83.45 | p = 0.30 (b = 0.35) |
|  | Insecticides | 12 | 7,456 | 1.43 | 1.00-2.05 | 0.0524 | 87.75 | p = 0.30 (b = -0.12) |
|  | Herbicides | 11 | 7,761 | **1.58** | **1.26-1.97** | **< 0.0001** | 72.49 | p = 0.06 (b = -0.03) |
|  | Fungicides | 4 | 2,649 | 1.45 | 0.70-2.99 | 0.3166 | 78.56 | p = 0.50 (b = -0.33) |
| Occupational | Any pesticides | 40 | 104,532 | **1.46** | **1.27-1.68** | **< 0.0001** | 71.37 | p = 0.15 (b = 0.11) |
|  | Insecticides | 20 | 237,515 | **1.46** | **1.13-1.88** | **0.0033** | 75.36 | p = 0.13 (b = -0.10) |
|  | Herbicides | 21 | 239,157 | **1.27** | **1.08-1.50** | **0.0045** | 59.37 | p = 0.005 (b = -0.19) |
|  | Fungicides | 15 | 236,158 | 1.25 | 0.99-1.60 | 0.0656 | 64.71 | p = 0.19 (b = -0.06) |
| Household | Any pesticides | 23 | 255,056 | **1.21** | **1.06-1.39** | **0.0058** | 75.15 | p = 0.87 (b = 0.16) |
|  | Insecticides | 8 | 4,209 | 1.29 | 0.98-1.70 | 0.0699 | 72.29 | p = 0.73 (b = 0.42) |
| Any type | Paraquat (herbicide) | 6 | 5,050 | **1.34** | **1.13-1.60** | **0.0007** | 49.92 | p = 0.98 (b = 0.28) |
| Occupational | Paraquat (herbicide) | 8 | 233,208 | 1.21 | 0.96-1.53 | 0.1061 | 72.31 | p = 0.053 (b = -0.27) |

Abbreviations: b, Egger's test intercept; CI, confidence interval; I<sup>2</sup>, Higgin's & Thompson's I<sup>2</sup> statistic; K, total number of studies on a pesticide exposure; n, total number of participants (counts) included in studies on a pesticide exposure; OR, odds ratio.
Bold text indicates statistically significant associations.

According to the meta-regression analyses for any pesticide class, insecticides, and herbicides, study quality, rural/urban setting, number of participants, study global subregion, method of exposure estimate, and the publication year did not influence heterogeneity, except for the rural/urban setting in any pesticide class and global subregion in herbicides. However, lower study quality, older published studies, small sample sizes, and global subregions had higher effect sizes in heterogeneity for fungicides.

### 3.3 Association between occupational pesticide exposure and Parkinson’s disease

Considering only occupational pesticide exposure, the meta-analysis showed that the use of any pesticide class (n = 40 studies) during lifetime increased the association with PD (OR = 1.46;

95% CI = 1.27 to 1.68) (Table 2), with high heterogeneity among studies (I^2^ statistics = 71.3%). For insecticides (n = 20) and herbicides (n = 21), the meta-analysis showed that exposure to these pesticide classes during the lifetime increased the association with PD (insecticides: OR = 1.46; 95% CI = 1.13 to 1.88; herbicides: OR = 1.27; 95% CI = 1.08 to 1.50) (Table 2), with high and moderate heterogeneity, respectively (insecticides: I^2^ statistics = 75.3%; herbicides: I^2^ statistics = 59.3%). For fungicides (n = 15), the meta-analyses showed a trend toward statistical significance for the association with PD (OR = 1.25; 95% CI = 0.99 to 1.60, p = 0.0656) (Table 2) with high heterogeneity (I^2^ statistics = 64.7%). According to Egger’s test, a publication bias was found among studies on occupational herbicide exposure.

According to the meta-regression analyses for any pesticide class, study quality, rural/urban setting, number of participants, study global subregion, method of exposure estimate, and publication year did not influence heterogeneity. However, the number of participants and publication year had some effect on heterogeneity for studies on insecticides and herbicides.

### 3.4 Association between household pesticide exposure and Parkinson’s disease

Considering only household pesticide exposure, the meta-analysis showed that the use of any pesticide class (n = 23 studies) during lifetime increased the association with PD (OR = 1.21; 95% CI = 1.06 to 1.39) (Table 2), with high heterogeneity among studies (I^2^ statistics = 75.1%). For insecticides (n = 8; number of participants = 4,209), the meta-analysis showed a trend toward statistical significance for the association with PD (OR = 1.29; 95% CI = 0.98 to 1.70, p = 0.0699) (Table 2), with high heterogeneity (I^2^ statistics = 72.2%). The limited number of studies on household pesticide exposure involving herbicides and fungicides did not permit meta-analyses. According to Egger’s test, no publication bias was detected among the studies.

According to the meta-regression analyses for any pesticide class or insecticides, study quality, rural/urban setting, number of participants, study global subregion, method of exposure estimate, and publication year did not influence heterogeneity.

### 3.5 Association between paraquat exposure and Parkinson’s disease

Considering only the use of paraquat, the meta-analysis showed that, when occupational and household use was not differentiated (n = 6 studies; number of participants = 5,050), the exposure to the product during lifetime increased the association with PD (OR = 1.34; 95% CI = 1.13 to 1.60) (Table 2), with moderate heterogeneity among studies (I^2^ statistics = 49.9%). For a strict occupational paraquat exposure (n = 8; number of participants = 233,208), the meta-analysis showed no association with PD (OR = 1.21; 95% CI = 0.96 to 1.53) (Table 2) with high heterogeneity (I^2^ statistics = 72.3%). The small number of studies on household paraquat exposure did not allow meta-analyses. According to Egger’s test, no publication bias was detected among the studies.

According to the meta-regression analyses for any type of paraquat exposure, study quality and global subregions were found to affect heterogeneity. For occupational paraquat exposure, the number of participants and the global subregion influenced the heterogeneity.

### 3.6 Sensitivity analyses

According to leading research groups, after excluding studies from the Ritz group (USA, n = 18), the associations between any household exposure and any paraquat exposure with PD were not significant (any household exposure: p = 0.2542; any paraquat exposure: p = 0.7127). Excluding studies from the Elbaz group (France, n = 7), no relevant association modifications were observed. According to study quality, analyses were impaired due to a low number of high-quality studies (NOS ≥ 6). Most associations remained unchanged for studies with poor or fair quality, as well as those with high quality.

## 4 DISCUSSION

The present systematic review provides additional evidence that any pesticide exposure during the lifetime, regardless of the type of exposure, may be associated with an increase in the association with PD by approximately 80%. This effect is particularly relevant for herbicides. Both specific occupational and household exposure are related to this effect. The herbicide paraquat is the product with the most substantial evidence of association with PD. Studies exhibit high methodological heterogeneity, primarily due to variations in pesticide measurement and analysis. These results align with previous systematic reviews and meta-analyses on the impact of pesticide exposure and the risk of developing PD ^4–14^.

Occupational exposure was the most extensively explored aspect of pesticide use in our systematic review, likely due to its agricultural applications. These products may affect nearly one billion rural workers worldwide ^18^, and the frequent health monitoring of these exposed populations living in rural regions with intense agricultural activity is justified. It may explain the high number of studies from two research groups in the USA and France, which analyzed rural counties in the Central Valley of California and individuals affiliated with the French *Mutualité Sociale Agricole*, respectively. According to our sensitivity analyses, the potential overrepresentation of studies from these rural settings in our meta-analyses affected the association with PD.

However, no previous systematic review analyzed the overall association between household pesticide exposure and PD. It is worth noting that nearly 4.4 billion people live in cities globally, and around 50 to 90% of urban individuals have reported using pesticides during their lifetime ^19^. Household pesticide use may pose a risk to human health, particularly in low- and middle-income countries, due to improper handling without the use of personal protective equipment and an informal market ^20^. According to our results, household pesticide exposure has a similar magnitude of association with PD to occupational exposure, suggesting that further studies on this type of exposure are needed ^21^.

Herbicides were the pesticide class most associated with PD in studies involving any type of exposure, particularly occupational exposure, with a high number of participants (n > 200,000). Household herbicide exposure description was rare. Previous meta-analyses have shown divergent results for herbicides ^6,9^. For occupational exposure, herbicides are the world’s most widely used pesticide class in agriculture ^22^. An earlier study performed a toxicogenomic analysis and found an association between PD and a list of 70 chemical products, including 14 herbicides ^23^. One of these herbicides is glyphosate, the most widely used herbicide worldwide ^24^.

Only occupational exposure to insecticides had a significant association with PD. Regarding household insecticide exposure, only a trend of association with PD was seen, possibly due to the low total number of participants. Previous meta-analyses have reported a positive association between any type of insecticide exposure ^6^, but not with occupational use ^6,9^. Also, there was no association between fungicide exposure and PD in our study and previous meta-analyses ^6,9^.

The low number of studies with household and specific pesticide class exposure may be due to data being mainly based on environmental questionnaires with high heterogeneity. Analyses that do not discriminate between the types of pesticide use or distinct exposure to herbicides, insecticides, and fungicides may impair the interpretation of results ^15^. Considering this, area-based geocoding techniques for estimating pesticide exposure may be viable alternatives to questionnaire-based studies, especially in rural areas with well-characterized land-use data and pesticide use reports ^25^.

Despite the high number of studies, there is a notable lack of diversity in the countries represented in these analyses. There is an overrepresentation of people from the USA, as approximately 40% of all included studies were from this country (with a total of 328,408 participants from the USA). In our systematic review, the two most populous countries, India and China, were represented by only four and two studies, respectively. These six studies recruited a total of fewer than 2,000 participants. Furthermore, the populations of the other top 10 most populous countries were not represented in the present study, including Indonesia, Pakistan, Nigeria, Bangladesh, and the Russian Federation.

The participation of some global regions, such as Latin America and the Caribbean, Sub-Saharan Africa, the Middle East and North Africa, Eastern Europe, Central Asia, and East and Southeast Asia, is markedly underrepresented. A similar pattern of underrepresentation of people from the Global South in PD research has been previously described in genetic studies ^26^. Considering occupational pesticide exposure, agricultural production is the main economic activity of many of these underrepresented countries. Additionally, the legal regulation of pesticide use in agriculture in low- and middle-income countries may be more permissive, allowing products with health issues ^27^.

This study has limited generalizability due to the lack of studies from already cited underrepresented regions and the low number of studies that categorized the type of exposure and specific pesticide class. Even though the results of the present study strengthen evidence for the association between pesticide exposure and PD. Also, the definition of the type of pesticide exposure and pesticide classes based on the authors’ description may not be precise, and this categorization was not evident in the text of some studies. However, for studies without a clear definition of the type of pesticide exposure and pesticide classes, we used broad categorizations (“any exposure” and “any class”).

Thus, previous meta-analyses and the present study indicate the same level of association between pesticide exposure and PD. However, we have to question the significance and relevance of these results in light of the methodological flaws in the original studies.

First, these studies were strongly based on data from environmental questionnaires for lifetime pesticide exposure, which are very heterogeneous among studies and subject to inaccuracy and recall bias, a form of differential misclassification bias influenced by time interval since exposure, personal characteristics of participants (age, education, socioeconomic status), the significance of the past event for individuals, social desirability of exposure, interviewing technique, and differential recall between cases and controls ^28^.

Second, excluding two cohort studies and seven cross-sectional studies (measurement of organochlorines in the blood at evaluation), all other 115 studies had a case-control design (where PD and control status served as the study inclusion criteria, and a retrospective analysis was conducted to determine which individuals were exposed to pesticides in the past) ^29^. As is known, observational retrospective studies cannot be used to establish a causal link between exposure and outcome, but may infer an association or correlation between them ^30,31^.

After dozens of published works on the theme since 1989, we can now argue that all evidence of an association between pesticide exposure and PD that could be inferred from this classic study design (case-control studies with environmental questionnaires) has already been extracted. But how do we increase the evidence of causality?

Recently, a review examined the causal link between various environmental risk factors and PD, including exposure to pesticides. According to the Bradford Hill criteria, the authors proposed that there is good evidence for an association between general pesticide exposure and PD in seven of the nine criteria. The authors proposed actions for three strategies (epidemiological, pathophysiological studies, and policy changes) to fill evidence gaps, including new studies with biomarkers for pesticide exposure ^27^. However, the development of new biomarkers is necessary due to the current lack of methods for measuring long-term exposure to most pesticides ^21^.

In conclusion, this systematic review updated the evidence that pesticide exposure is a risk factor for developing PD, regardless of the type of exposure. The association may depend on the pesticide class, being stronger for herbicides. Because results from classic case-control studies with environmental questionnaires are limited in explaining causality, further studies with new methods of pesticide exposure measurement and innovative design studies in underrepresented populations are still needed.

## Funding

P.H.P.S., J.S.A., R.S.G.G., and H.K.A.O. report no disclosures. A.F.S.S. has received grants from the Michael J. Fox Foundation (MJFF), the Aligning Science Across Parkinson’s Global Parkinson Genetic Program (ASAP-GP2), Brazilian National Council for Scientific and Technological Development (CNPq - Brazil), Fapergs (Brazil), and honoraria from MDS. I.F.M. has received grants from the Stanley Fahn Junior Faculty Award and an International Research Grants Program award both from the Parkinson’s Foundation and by a research grant from the American Parkinson’s Disease Association. He is currently funded by the National Institutes of Health, MJFF, and the ASAP-GP2. B.L.S-L. has received a grant from the CNPq - Brazil.

## Data Availability

The data that supports the findings of this study are available in an online public repository

## Notes

**Conflict of interest disclosure:** Nothing to report

### Competing Interest Statement

The authors have declared no competing interest.

### Clinical Protocols

https://www.crd.york.ac.uk/prospero/

### Funding Statement

This study did not receive any funding

